# Clinical Characteristics and Outcomes of Venous Thromboembolism in Patients Hospitalized for COVID-19: Systematic Review and Meta-Analysis

**DOI:** 10.1101/2020.06.14.20130922

**Authors:** Joshua Henrina, Iwan Cahyo Santosa Putra, Irvan Cahyadi, Hoo Felicia Hadi Gunawan, Alius Cahyadi, Leonardo Paskah Suciadi

## Abstract

**Objective:** To investigate the clinical characteristics and outcomes of Coronavirus Disease of 2019 (COVID-19) patients complicated with venous thromboembolism (VTE)

**Method:** We performed a comprehensive literature search of several databases to find studies that assessed VTE in hospitalized COVID-19 patients with a primary outcome of all-cause mortality and secondary outcomes of intensive care unit (ICU) admission and mechanical ventilation. We also evaluated the clinical characteristics of VTE sufferers.

**Results:** Eight studies have been included with a total of 1237 pooled subjects. Venous thromboembolism was associated with higher mortality (RR 2.48 (1.35, 4.55), *p*=0.003; *I*^*2*^ 5%, *p=*0.35) after we performed sensitivity analysis, ICU admission (RR 2.32 (1.53, 3.52), *p*<0.0001; *I*^*2*^ 80%, *p* <0.0001), and mechanical ventilation need (RR 2.73 (1.56, 4.78), *p=*0.0004; *I*^*2*^ 77%, *p=*0.001). Furthermore, it was also associated to male gender (RR 1.21 (1.08, 1.35), *p*=0.0007; *I*^*2*^ 12%, *p*=0.34), higher white blood cells count (MD 1.24 (0.08, 2.41), 0.04; *I*^*2*^ 0%; 0.26), D-dimer (MD 4.49 (2.74, 6.25), *p*<0.00001; *I*^*2*^ 67%, *p*=0.009) and LDH levels (MD 70.93 (19.33, 122.54), *p<*0.007; *I*^*2*^ 21%, p=0.28). In addition, after sensitivity analysis was conducted, VTE also associated with older age (MD 2.79 (0.06, 5.53), *p*=0.05; *I*^*2*^ 25%, *p=*0.24) and higher CRP levels (MD 2.57 (0.88, 4.26); *p=*0.003; *I*^*2*^ 0%, *p*=0.96).

**Conclusion:** Venous thromboembolism in COVID-19 patients was associated with increased mortality, ICU admission, and mechanical ventilation requirement. Male gender, older age, higher levels of biomarkers, including WBC count, D-Dimer, and LDH were also being considerably risks for developing VTE in COVID-19 patients during hospitalization.

## Introduction

Coronavirus disease of 2019 (COVID-19) is a highly contagious respiratory disease caused by Severe Acute Respiratory Syndrome Coronavirus type 2 (SARS-CoV-2) resulting in a global pandemic status. As of this paper writing, it has already claimed more than 400.000 lives and infected approximately 7 million people throughout the world (1). As the name implies, many aspects of this multifaceted disease are being explored. Notably, the coagulopathy associated with COVID-19, in which the evidence is still emerging.(2–5)

A previous study has shown that D-dimer, a primary marker of coagulopathy, was associated with higher mortality.(5) Furthermore, lung dissection and autopsy from COVID- 19 decedents showed pulmonary microthrombi formation.(4,6) Therefore, several authors argued that Acute Respiratory Distress Syndrome (ARDS), which is the most common complication of COVID-19, is caused by pulmonary endothelial dysfunction identified as pulmonary intravascular coagulopathy (PIC) or pulmonary thrombosis.(7,8) This argument is substantiated by several findings that the incidence of pulmonary embolism (PE) is more frequent than deep vein thrombosis (DVT).(9)

Data from multiple studies have shown a higher incidence of venous thromboembolism (VTE) in hospitalized COVID-19 patients compared to other diseases.(10,11) This acute vascular complication might potentially affect morbidity and mortality of COVID-19 patients during hospitalization. Nevertheless, a retrospective study conducted by Tang et al. showed that patients with higher D-dimer levels and sepsis-induced coagulopathy (SIC) scores who received heparin have lower mortality rates.(4) However, more prospective studies are needed to confirm this result.

Therefore, the study aimed to systematically investigate the outcomes of hospitalized COVID-19 patients complicated with VTE. We also scrutinized any clinical characteristics which potentially indicated to developing VTE in this population during hospitalization. With this study, we want to determine which COVID-19 patients that have the propensity to develop VTE during hospitalization. Thus, more attention can be given to these patients to reduce morbidity and mortality during hospitalization.

## Methods

### Search strategy

The predefined protocol was used for this review and in line with Preferred Reporting Items for Systematic Reviews and Meta-Analysis (PRISMA). We systematically searched and identified studies published between 1 January 2020 and 31 May 2020, through electronic searches using MEDLINE, Europe PMC, Proquest, Ebscohost, Google Scholar, Cochrane Central, Clinicaltrial.gov and from preprint websites (Preprint.org and MedRxiV.org). We used Medical Subject Headings (MeSH) and free word to construct our search terms related to venous thromboembolism (venous thromboembolism OR deep vein thrombosis OR pulmonary embolism), coronavirus disease of 2019 (COVID-19 and SARS-CoV-2). We identified articles eligible for further review by performing screening of identified titles or abstracts, followed by a full-text review. In the case of multiple publications, the most recent and complete report was included. We finalized the systematic literature review search on 31 May 2020. This study adhered to the PRISMA guideline.

### Selection criteria

Articles eligibility was assessed by two independent investigators (JH and ICSP). Discrepancies were resolved by consensus with a third investigator (AC). The inclusion criteria of the article in this study included all research studies involving hospitalized COVID-19 patients experiencing VTE (DVT group and PE group only, non-specified VTE group, and specified VTE group). We excluded commentaries, non-research letter, reviews, and case reports/case series.

### Data extraction

A standardized form was used to collect the information that consists of qualitative aspects of identified studies (the first author and publication year, geographic locations, and study design), COVID-19 patients’ profile (gender and age), venous thromboembolisms, and comorbidities. Two authors performed data extraction independently (JH and ICSP). The primary outcome of interest was all-cause mortality, and the secondary outcomes of interest were intensive care admission and mechanical ventilation. In addition, we also extracted data regarding clinical characteristics (patient characteristics, comorbidities, sign and symptoms, and laboratory findings). Data that was reported other than mean ± SD was transformed accordingly using the formula created by Wan et al.(12)

### Statistical analysis

We performed meta-analysis using Review Manager 5.4 (https://training.cochrane.org/online-learning/core-software-cochrane-reviews/revman). For dichotomous variables, we calculated the pooled estimates and its 95% confidence interval in the form of risk ratios (RRs) using the Mantel-Haenzel formula. We calculated the pooled estimates for continuous variables in the form of a mean difference (MD) and its standard deviation. To account for interstudy variability regardless of the heterogeneity, we used a random-effects model. We used two-tailed p values with a significance set at ⍰0.05. To assess heterogeneity across studies, we used the inconsistency index (I^2^) with a value above 50% or p < 0.05 indicates significant heterogeneity. We performed a sensitivity analysis using the leave-one-out method to achieve statistical robustness and find the source of heterogeneity. Finally, to qualitatively detect publication bias, we used an inverted funnel-plot analysis.

## Results

### Study selection and characteristics

Initial systematic database search yielded 365 studies, of which 192 remain after duplicates were excluded. Through title/abstract screening, 170 studies were excluded and left with 22 remaining studies. The final 22 studies were assessed for full-text eligibility, of which 15 of them were excluded due to: one was editorial, two were case reports, three were case-series, one study of PE in non-COVID-19 patients, one study in oupatient COVID-19 patients, and six studies in ICU admitted COVID-19. As a result, only eight studies (n= 1237 subjects) were included in the qualitative and quantitative synthesis.(13–19) (Figure 1, Table 1)

**Figure 1.**
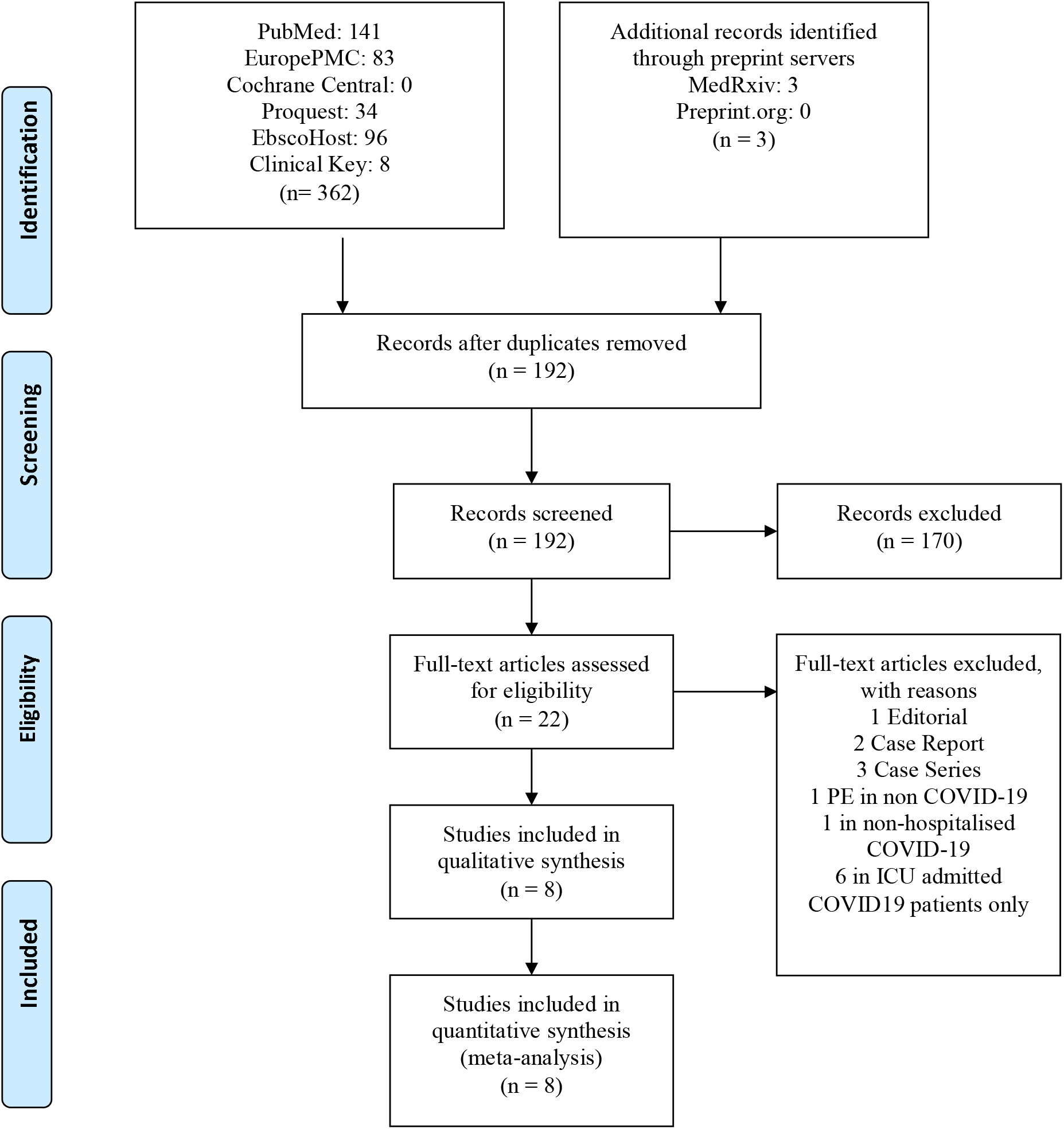
Preferred Reporting Items for Systematic Reviews and Meta-Analyses (PRISMA) study flow diagram. COVID-19, Coronavirus disease of 2019.

**Table 1.** Characteristics of included studies

| Author | Country | Study design | Sample size | VTE/PE/DVT (%) | Age (mean±SD) | Male (%) | Hypertension (%) | Diabetes Mellitus (%) | Smoking (%) | COPD/ respiratory disease (%) | Primary Outcome | Secondary Outcome |
| --- | --- | --- | --- | --- | --- | --- | --- | --- | --- | --- | --- | --- |
| Mathieu Artifoni | France | Retrospective cohort | 71 | VTE; 16 (22,54) | 60.2 ± 36.5<br>vs<br>62.2 ± 30.6 | 11/16 (68.7)<br>vs<br>32/55 (58.2) | 3/16 (19)<br>vs<br>26/55 (47) | 0/16 (0)<br>vs<br>14/55 (25) | 0/16 (0)<br>vs<br>6/55 (12) | NA | Mortality | ICU admission<br>Mechanical ventilation |
| Saskia Middeldorp | Netherlands | Retrospective cohort | 198 | VTE; 39 (19,7) | 62 ± 10<br>vs<br>60 ± 15 | 27/39 (69)<br>vs<br>103/159 (65) | NA | NA | NA | NA | NA | ICU admission |
| Ian Leonard Lorant | France | Retrospective cohort | 106 | PE; 32 (30,19) | 64 ± 22<br>vs<br>63 ± 15 | 25/32 (78)<br>vs<br>45/74 (57) | NA | NA | NA | NA | NA | ICU admission |
| Neo Poyiadji | USA | Retrospective cohort | 328 | PE; 72 (21,95) | 59 ± 15<br>vs<br>62 ± 16 | 35/72 (49)<br>vs<br>115/256 (45) | 41/72 (57)<br>vs<br>156/256 (61) | 29/72 (40)<br>vs<br>97/256 (38) | 20/72 (28)<br>vs<br>33/256 (13) | 5/72 (7)<br>vs<br>102/256(40) | Mortality | ICU admission<br>Mechanical ventilation |
| Franck Grillet | France | Retrospective cohort | 100 | PE; 23 (23) | 67 ± 11<br>vs<br>66 ± 13 | 21/23 (91)<br>vs<br>49/77 (64) | NA | 6/23 (23)<br>vs<br>14/77(18) | NA | 4/23 (17)<br>vs<br>10/77 (13) | NA | ICU admission |
| Florian Bompard | France | Retrospective cohort | 135 | PE; 32 (23,7) | 68.6 ± 20<br>vs<br>63.3 ± 25.27 | 26/32 (81)<br>vs<br>68/103 (66) | NA | NA | NA | NA | NA | ICU admission<br>Mechanical ventilation |
| P. Demelo-Rodríguez | Spain | Prospective cohort | 156 | DVT; 23 (14,7) | 66.7 ± 15.2<br>vs<br>68.4 ± 14.4 | 14/23 (60.9)<br>vs<br>88/133 (66.2) | NA | NA | NA | NA | NA | ICU admission |
| Liming Zhang | China | Cross Sectional | 143 | DVT; 66 (46.1) | 67 ± 12<br>vs<br>59 ± 16 | 36/66 (54.5)<br>vs<br>38/77 (49.4) | 28/66 (42.4)<br>vs<br>28/77 (36.4) | 13/66 (19.7)<br>vs<br>13/77 (16.9) | 5/66 (7.6)<br>vs<br>4/77 (5.2) | 5/66 (7.6)<br>vs<br>4/77 (5.2) | Mortality | ICU admission<br>Mechanical ventilation |

a. VTE; venous thromboembolism, PE; pulmonary embolism, DVT; deep vein thrombosis, COPD; chronic obstructive pulmonary disease, NA; not available

### Venous thromboembolism and primary outcome

The initial meta-analysis showed that complication of VTE in COVID-19 patients was not associated with higher mortality (RR1.63 (0.63, 4.24); *p*=0.31; *I*^*2*^ 64%, *p*= 0.04). However, the result was significant after sensitivity analysis was conducted, by excluding one study (Poyiadji et al.); (RR 2.48 (1.35, 4.55), *p=*0.003; *I*^*2*^ 5%, *p=*0.35). (Table 2, Figure 2 and 3)

**Table 2.** Summary of meta-analysis of primary and secondary outcomes

| Variable | Risk Ratio (95%CI);<br>*p*-value | Heterogeneity<br>(*I*<sup>2</sup>, *p*-value) | Number of Studies |
| --- | --- | --- | --- |
| *Primary Outcome* |  |  |  |
| Mortality | 1.63 (0.63, 4.24); 0.31 | 64%; 0.04 | 4 |
| Mortality SA<sup>a</sup> | 2.48 (1.35, 4.55); 0.003 | 5%; 0.35 | 3 |
| *Secondary Outcomes* |  |  |  |
| ICU Admission | 2.32 (1.53, 3.52); <0.0001 | 80%; <0.0001 | 7 |
| ICU Admission SA<sup>a</sup> | 2.09 (1.38, 3.16); 0.0005 | 70%; 0.003 | 6 |
| Mechanical Ventilation | 2.73 (1.56, 4.78); 0.0004 | 77%; 0.001 | 4 |
| Mechanical Ventilation SA | 3.35 (2.48, 4.53); <0.00001 | 17%; 0.30 | 3 |

a. CI; confidence interval, SA; sensitivity analysis by leave-one-out method

**Figure 2.**
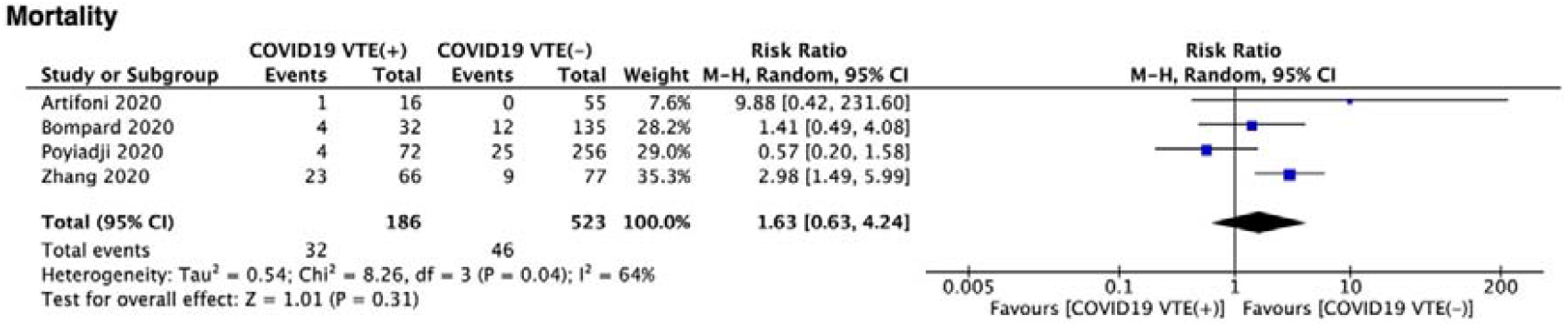
Venous thromboembolism (VTE) and mortality. There was no difference in all-cause mortality between the VTE group and the non- VTE group.

**Figure 3.**
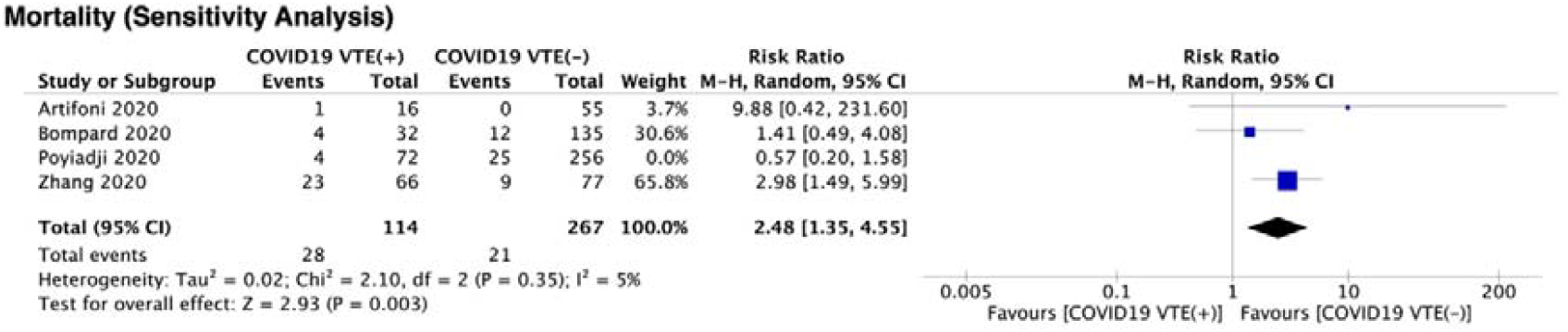
Venous thromboembolism (VTE) and mortality. After excluding one study (Poyiadji et al.), the COVID-19 with VTE was associated with higher all-cause mortality.

### Venous thromboembolism and secondary outcome

Venous thromboembolism in patients with COVID-19 was significantly associated with higher ICU admission (RR 2.32 (1.53, 3.52), *p*<0.0001; *I*^2^ 80%, *p* <0.0001). This result is still consistent after performing sensitivity analysis by excluding study by Middeldorp et al., in addition to lower heterogeneity (RR 2.09 (1.38, 3.16), *p*=0.0005; *I*^2^70%, *p*=0.003). (Table 2, figure 4 and 5) Furthermore, VTE occurrence indicated the more need for mechanical ventilation support during hospital stay (RR 2.73 (1.56, 4.78), *p=*0.0004; *I*^2^ 77%, *p=*0.001). A similar expected result, along with reduced heterogeneity, was noted after a sensitivity analysis was performed, excluding study by Poyiadji et al. (RR 3.35 (2.48, 4.53), *p*<0.00001; *I*^2^ 17%, *p*= 0.30). (Table 2, figure 6 and 7)

**Figure 4.**
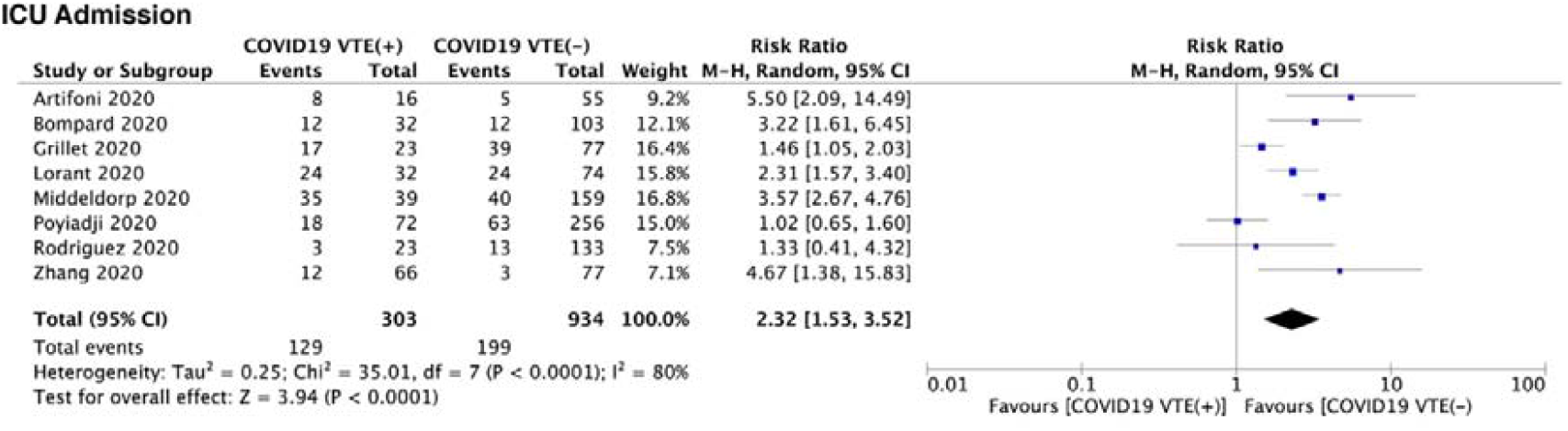
Venous thromboembolism and ICU admission. ICU admission is higher in COVID-19 patients with VTE compared to the non-VTE group.

**Figure 5.**
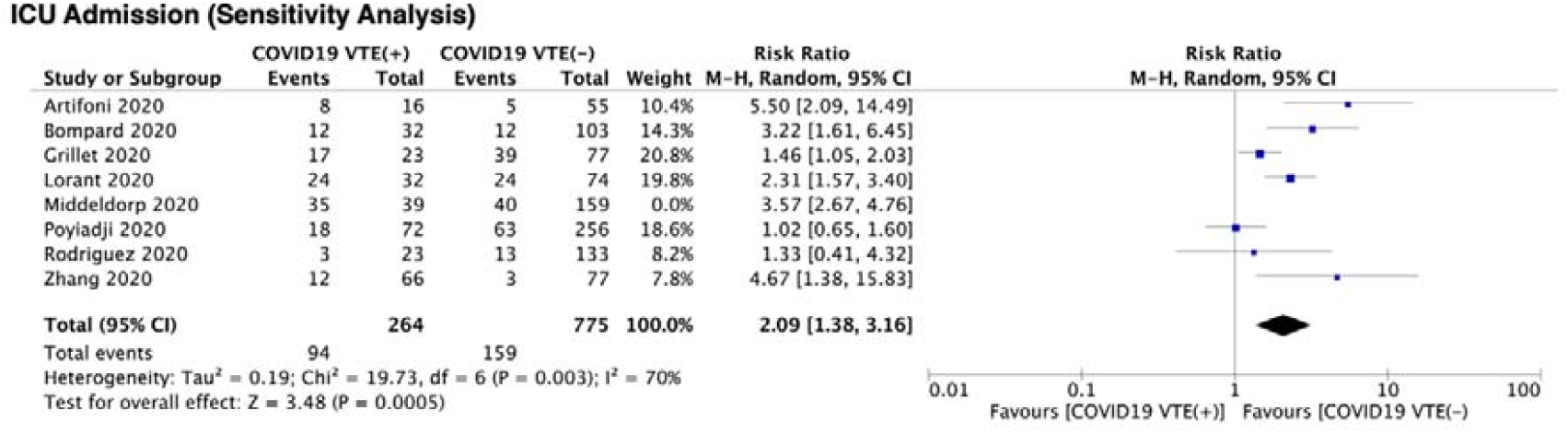
Venous thromboembolism and ICU admission. After excluding one study (Middeldorp et al.), the heterogeneity can be reduced while maintaining significance.

**Figure 6.**
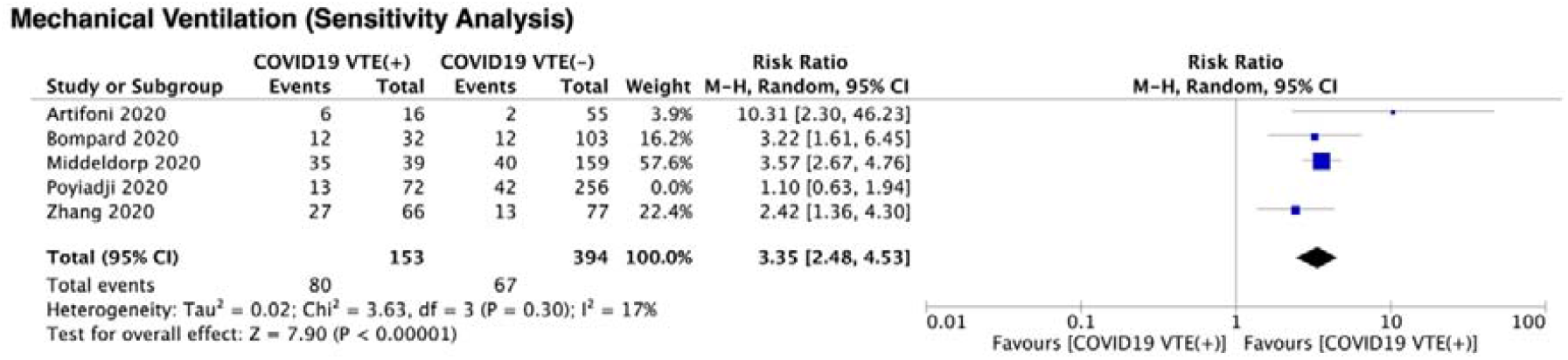
Venous thromboembolism and mechanical ventilation. COVID-19 patients complicated with VTE were associated with higher need of mechanical ventilation.

**Figure 7.**
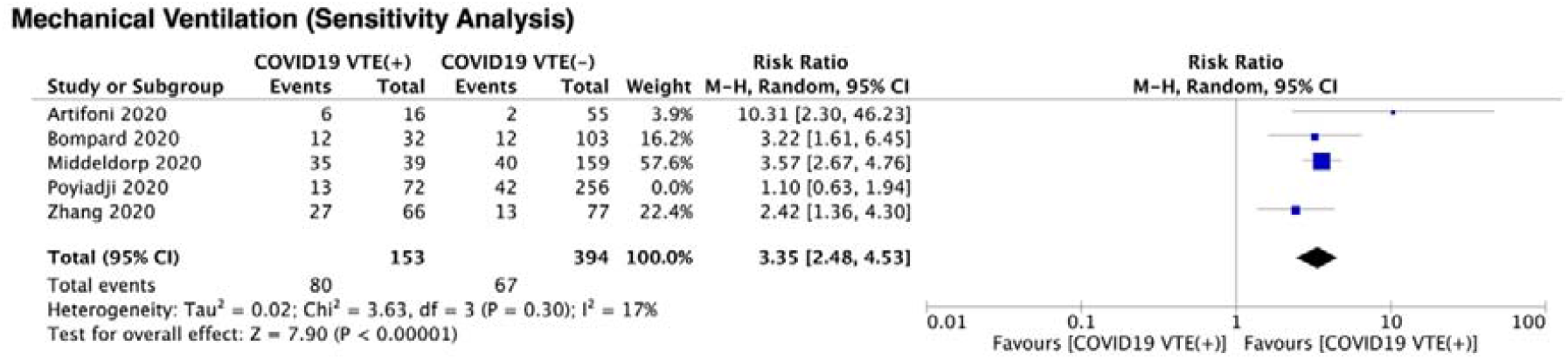
Venous thromboembolism and mechanical ventilation. After excluding one study (Poyiadji et al.), the heterogeneity can be reduced while maintaining significance.

### Clinical characteristics associated with VTE

There were 27 variables consist of patient characteristics, comorbidities, VTE risk factors, signs and symptoms, and laboratory findings have been analysed (Table 3 and Table 4). However, there were only 8 variables associated with VTE, and three of them only included two studies.

**Table 3.**
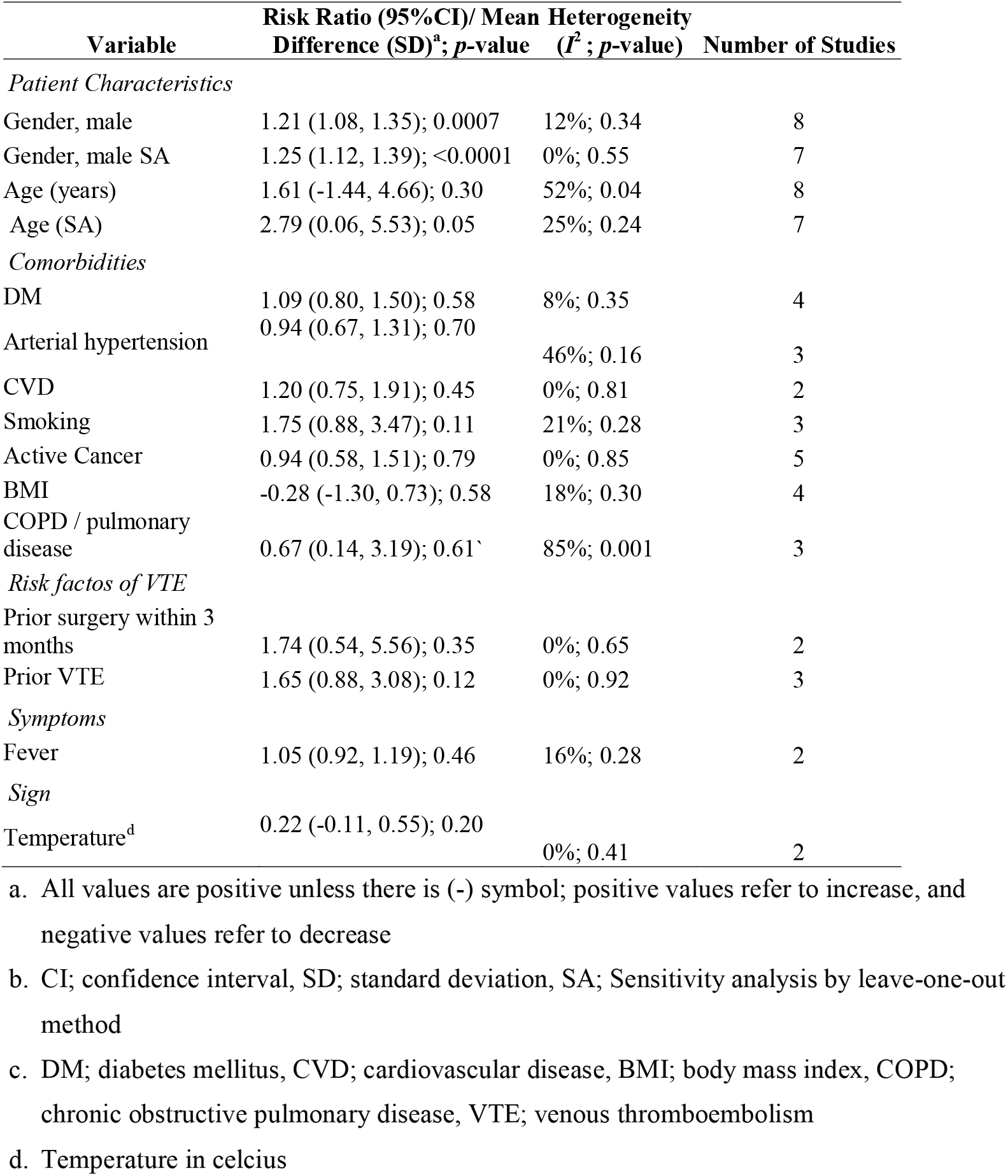
Summary of meta-analysis of demographic profiles, comorbidities, and clinical manifestations.

| Variable | Risk Ratio (95%CI)/ Mean<br>Difference (SD)<sup>a</sup>; *p*-value | Heterogeneity<br>(*I*<sup>2</sup>; *p*-value) | Number of Studies |
| --- | --- | --- | --- |
| *Patient Characteristics* |  |  |  |
| Gender, male | 1.21 (1.08, 1.35); 0.0007 | 12%; 0.34 | 8 |
| Gender, male SA | 1.25 (1.12, 1.39); <0.0001 | 0%; 0.55 | 7 |
| Age (years) | 1.61 (-1.44, 4.66); 0.30 | 52%; 0.04 | 8 |
| Age (SA) | 2.79 (0.06, 5.53); 0.05 | 25%; 0.24 | 7 |
| *Comorbidities* |  |  |  |
| DM | 1.09 (0.80, 1.50); 0.58 | 8%; 0.35 | 4 |
| Arterial hypertension | 0.94 (0.67, 1.31); 0.70 | 46%; 0.16 | 3 |
| CVD | 1.20 (0.75, 1.91); 0.45 | 0%; 0.81 | 2 |
| Smoking | 1.75 (0.88, 3.47); 0.11 | 21%; 0.28 | 3 |
| Active Cancer | 0.94 (0.58, 1.51); 0.79 | 0%; 0.85 | 5 |
| BMI | -0.28 (-1.30, 0.73); 0.58 | 18%; 0.30 | 4 |
| COPD / pulmonary disease | 0.67 (0.14, 3.19); 0.61<sup>^</sup> | 85%; 0.001 | 3 |
| *Risk factors of VTE* |  |  |  |
| Prior surgery within 3 months | 1.74 (0.54, 5.56); 0.35 | 0%; 0.65 | 2 |
| Prior VTE | 1.65 (0.88, 3.08); 0.12 | 0%; 0.92 | 3 |
| *Symptoms* |  |  |  |
| Fever | 1.05 (0.92, 1.19); 0.46 | 16%; 0.28 | 2 |
| *Sign* |  |  |  |
| Temperature<sup>d</sup> | 0.22 (-0.11, 0.55); 0.20 | 0%; 0.41 | 2 |

- a. All values are positive unless there is (-) symbol; positive values refer to increase, and negative values refer to decrease - b. CI; confidence interval, SD; standard deviation, SA; Sensitivity analysis by leave-one-out method - c. DM; diabetes mellitus, CVD; cardiovascular disease, BMI; body mass index, COPD; chronic obstructive pulmonary disease, VTE; venous thromboembolism - d. Temperature in celcius

**Table 4.**
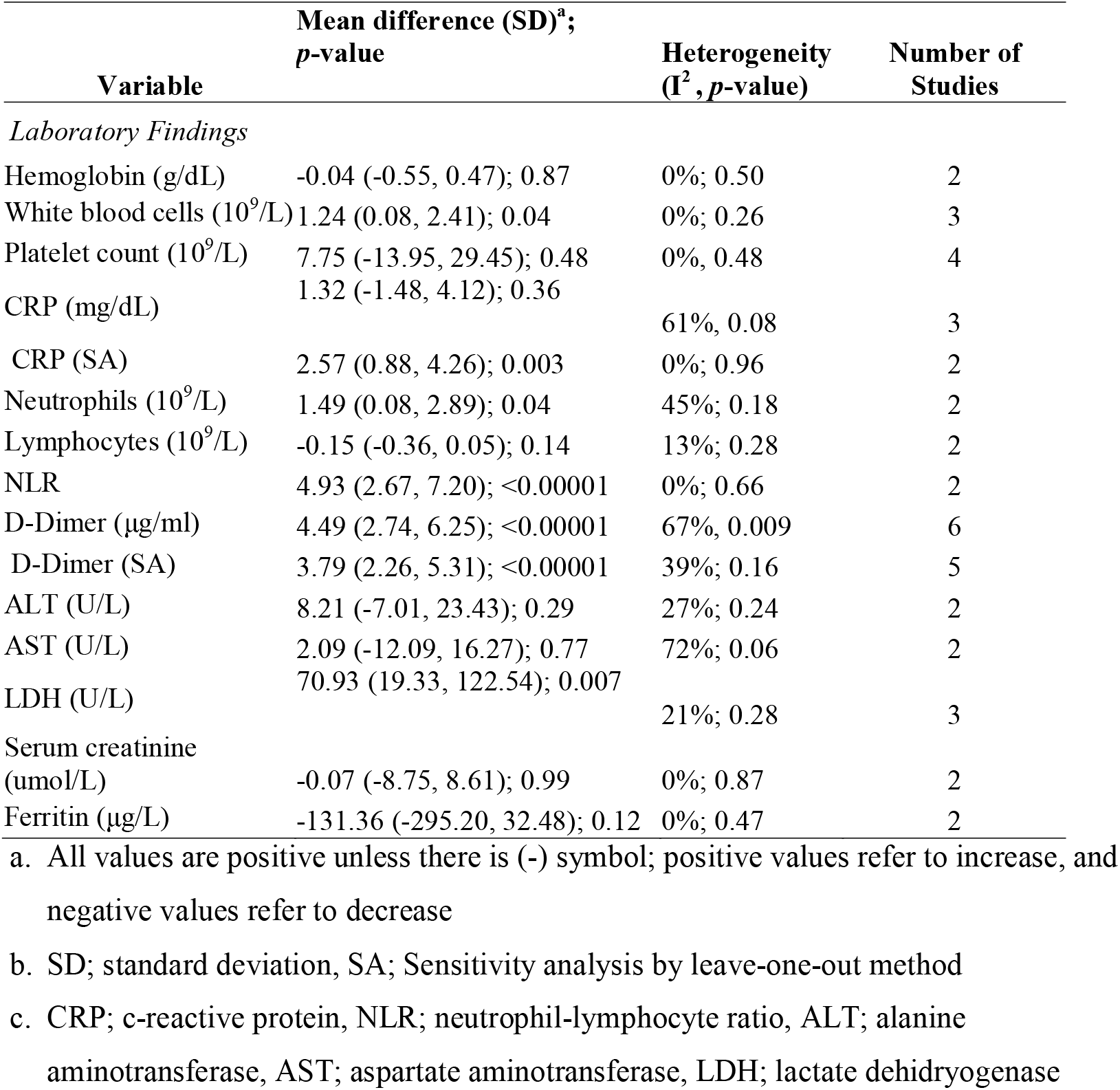
Summary of meta-analysis of laboratory findings

The meta-analysis showed that the incidence of VTE in COVID-19 patients were more common in men (RR 1.21 (1.08, 1.35), *p*=0.0007; *I*^*2*^ 12%, *p*=0.34). Sensitivity analysis by excluding one study (Rodriguez) showed that heterogeneity could be reduced while maintaining significance. (RR 1.25 (1.12, 1.39), *p*<0.0001; *I*^*2*^ 0%, *p*=0.55) Even though it was not associated with older age (MD 1.61 (-1.44, 4.66), *p* =0.30; *I*^*2*^ 52%, *p=*0.04), sensitivity analysis by excluding study by Poyiadji et al. redirected that older age contributed to increased risk for developing VTE during hospitalization (MD 2.79 (0.06, 5.53), *p*=0.05; *I*^*2*^ 25%, *p=*0.24).

Additionally, higher levels of white blood cell (WBC) count (MD 1.24 (0.08, 2.41), 0.04; *I*^*2*^ 0%; 0.26), D-dimer (MD 4.49 (2.74, 6.25), *p*<0.00001; *I*^*2*^ 67%, *p*=0.009) and LDH levels (MD 70.93 (19.33, 122.54), *p<*0.007; *I*^*2*^ 21%, p=0.28) were also associated with VTE complications in COVID-19. Sensitivity analysis on D-Dimer by excluding a study by Poyiadji et al. revealed that heterogeneity can be reduced, an indication of statistical robustness (RR 3.79 (2.26, 5.31), *p*<0.00001; *I*^*2*^ 39%, 0.16). Higher CRP levels also associated with VTE after we performed sensitivity analysis by excluding the Rodriguez et al. study (MD 2.57 (0.88, 4.26); *p<*0.003; *I*^*2*^ 0%, *p*=0.96). Other parameters including neutrophils count and neutrophil-lymphocyte ratio might be potentially associated with VTE in COVID-10 (Table 4). However, CRP levels and these variables were only available in two studies (Figure 7).

### Subgroup analysis

We only carried out subgroup analysis consisted of DVT only, PE only, and unspecified VTE group in correlation to ICU admission. Other subgroup analyses for mortality and mechanical ventilation were unable to be conducted due to two subgroups (DVT and VTE) contained less than two studies. As a result, the DVT only group was not associated with ICU admission (RR 2.47 (0.71, 8.59), *p=*0.16; *I*^*2*^ 54%, *p*=0.14). Whereas in contrast, higher ICU admission were significant in the PE only group (RR 1.75 (1.13, 2.69), *p=*0.01; *I*^*2*^ 74%, *p*=0.009) and in unspecified VTE group (RR 5.50 (2.09, 14.49), *p*<0.00001; *I*^*2*^ 0%, *p*=0.39). However, considerable heterogeneity between subgroups was noted (*I*^*2*^ 75.9%, *p*=0.02). (Figure 8)

**Figure 8.**
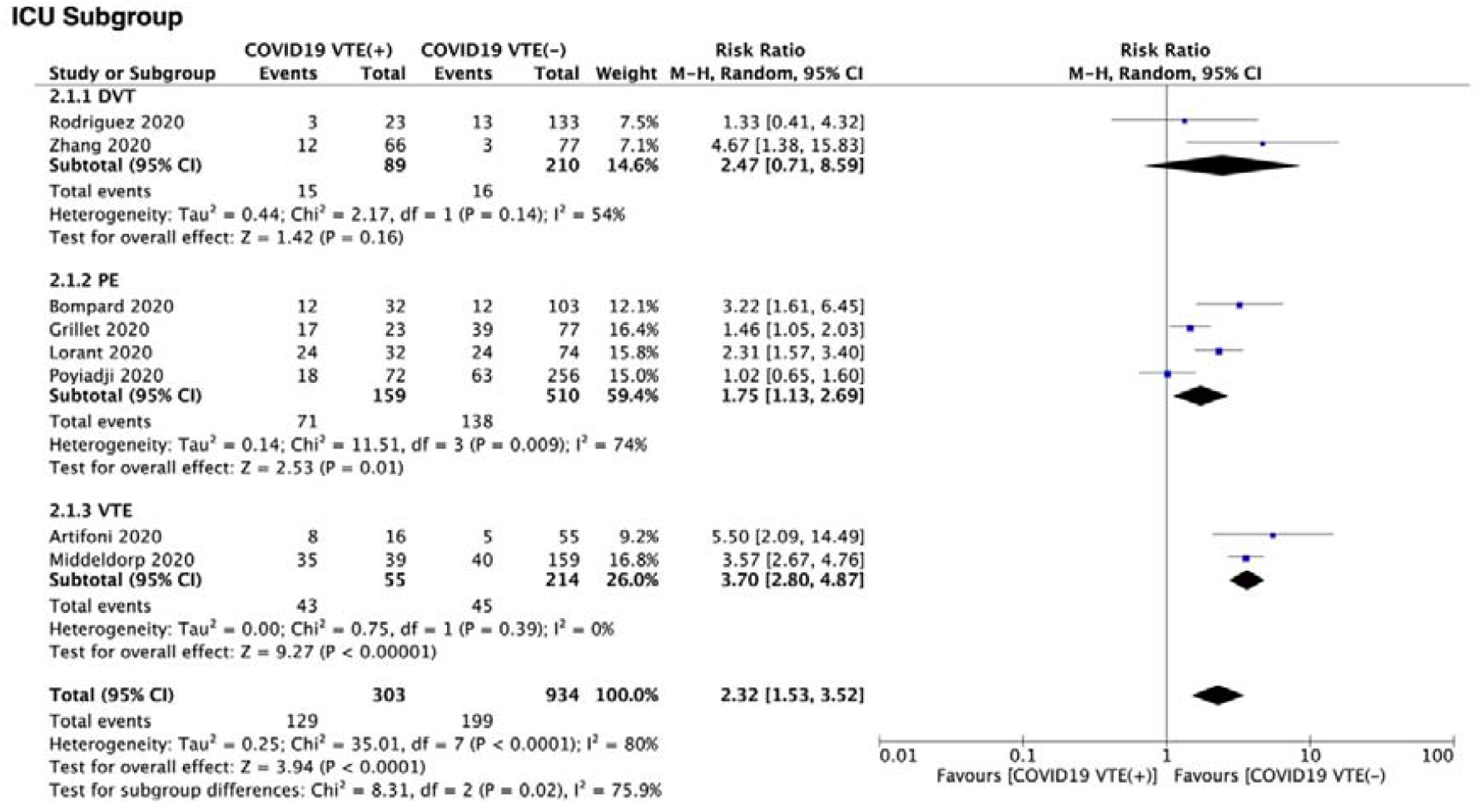
Subgroup analysis of venous thromboembolism and ICU admission. COVID-19 patients complicated with PE and VTE, were associated with higher ICU admission. However, considerable heterogenity and subgroup differences were noted.

### Publication bias

We did not perform publication bias in accordance to the Cochrane handbook reference which stipulates that the power of inverted funnel-plot analysis is too low to discriminate the chance from real asymmetry if the included studies are less than 10.(21)

## Discussions

Coagulopathy and VTE in COVID-19 are an emerging phenomenon with devastating consequences and has been associated with poor prognosis in COVID-19.(22,23) Thus, it prompts International Society of Thrombosis and Haemostasis (ISTH) to publish interim guidelines for managing COVID-19’s coagulopathy.(24) They recommend that all hospitalized COVID-19 patients, should be prophylactically anticoagulated.

The underlying factors contributed to VTE according Virchow’s triad, which are venous stasis, activation of coagulation, and vascular dysfunction.(25) The first factor, venous stasis, is common due to immobilization in hospitalized patients, especially patients that reside in the ICU. Nonetheless, the incidence of VTE in ICU-admitted COVID-19 patients were higher compared to other diseases.(10,11)

The other two factors are culmination of COVID-19 infection. SARS-CoV-2 can infect different organs through angiotensin-converting enzyme 2 (ACE-2), a glycoprotein metalloprotease that are ubiquitously found in human organs, particularly the endothelial cells, which traversed multiple organs and the pneumocytes.(26,27) The ensuing hypoxia caused by lung injury and ARDS in COVID-19 is a key factor of vascular fibrin deposition due to the increased early growth response-1 (Egr-1), which triggered tissue factor transcription/translation by mononuclear phagocytes and smooth muscle cells.(28) Furthermore, there is a role of bidirectional immunothrombosis, which is heightened inflammation will shift the balance to the procoagulant state by downregulating necessary anticoagulant pathways, while upregulating tissue factor expression necessary for thrombin generation.(29)

In addition, SARS-CoV-2 downregulates ACE-2 after successfully infected cells, which is a counterregulatory enzyme of Angiotensin II (AngII) that maintains the renin-angiotensin system (RAS) balance in check.(30) This condition leads to a prothrombotic milieu. Interaction of AngII with angiotensin II type 1 receptor (ATR-1) will activate transcription factors, which increased adhesion and accumulation of neutrophils and macrophages to endothelial cells, and ultimately these cells will release proinflammatory cytokines and chemokines.(30,31) All of these will preserve the vicious cycle of inflammation and coagulopathy.

SARS-CoV-2 is also known to cause direct vascular injury via ACE-2 receptors. Case series of post mortem analysis of decedents due to COVID-19 confirmed diffuse endothelial inflammation, termed endothelitis, viral elements within endothelial cells, and inflammatory cell death.(32) Furthermore, morphologic and molecular changes from autopsied decedents due to COVID-19 ARDS showed severe endothelial injury with viruses present intracellularly and disrupted cell membranes.(33) Another hallmark feature was thrombosis and microangiopathy and intussusceptive angiogenesis, which was nine times and 2.7 times more common than patients with influenza.(34)

To the best of our knowledge, this is the first systematic review and meta-analysis that describes the clinical characteristics and outcomes associated with VTE in COVID-19 patients. The meta-analysis has shown that VTE was significantly associated with mortality compared to the non-VTE group after sensitivity analysis, by excluding Poyiadji et al. Of note, higher all-cause mortality was seen in non VTE COVID-19 patients in this study.

This finding is in accordance with a study from China by Wang et al., involving 1026 subjects with laboratory-positive COVID-19 that showed a high risk of thromboembolism, defined as Padua score of ≥4, which is significantly associated with mortality.(35) Despite pooling mortality risk ratio across all VTE phenomena, which might introduced bias due to mortality disparity between PE only group and DVT only group, a study by Zhang et al. showed that COVID-19 patients with DVT had significant risks of developing adverse outcomes consist of more ICU admission (p=0.005), fewer hospital discharges (p<0.001), and more deaths (p=0.001).(18) Thus, it is justified to include DVT under the same umbrella term of VTE with PE. Furthermore, Wang et al. also showed that these patients were more likely to have been admitted to the ICU.(35) We also find this association to be true. In comparison to the non-VTE group, the VTE group was significantly associated with higher ICU admission. Also, our data showed that VTE patients were likely to be mechanically ventilated, confirming aforementioned study.(35)

However, it is worth mentioning, that one of the included studies highlighted that 72% pulmonary embolism cases was diagnosed in patients who do not need ICU level care.(15) In addition, Rodriguez et al. reported that asymptomatic DVT was diagnosed in 23 out of 156 patients (14.7%).(17)Thus, special attention is needed for asymptomatic or ambulatory COVID-19 patients.(22)

It is well known that worse clinical presentations of COVID-19 disproportionately affect the older population and male gender. However, the pathomechanisms remain elusive. In our meta-analysis, the male gender was associated with VTE. This result might explain, in part, why males were disproportionately affected. Nonetheless, men do have a higher risk of first and recurrent VTE compared to women, and it is hypothesized due to X or Y link mutation or mutation on a gene with a sex-specific effect.(36)

In our meta analysis, VTE was associated with the older population after we performed sensitivity analysis, by excluding Poyiadji.(15) In his study, VTE occured in younger patients. Younger populations in VTE sufferers were also noted in studies by Artifoni et al. and Rodriguez et al.(13,17) However, it is important to keep in mind that analysis from a population-based study of 25 years involving 2218 patients, with increasing age, there is a marked increase in VTE incidences in both sexes.(37)

Previously, a meta-analysis by Tian, et al. found that higher WBC count, D-Dimer, CRP, and LDH levels were associated with mortality in COVID-19.(38). These biomarkers serve as an indicator of heightened inflammation and organ damages. Thus, in the setting of marked inflammation, the bidirectional role of immunothrombosis plays a crucial part in VTE’s pathomechanism. In our meta-analysis, higher WBC count, D-Dimer, CRP, and LDH levels were associated with VTE. However, the CRP variable gains significance after we only included two studies.(table 4) Therefore, more prospective studies are needed to better understand the relevance of CRP levels as a risk marker.

On the other hand, our meta-analysis did not yield any association between VTE and comorbidities. Therefore, another likely explanation of why there is a higher burden of COVID-19 in patients with comorbidities.(30,39) Furthermore, it is not surprising that symptoms and signs other than dyspnea were not associated with VTE. Also, dyspnea in COVID-19 is not adequately sensitive to distinguish pulmonary embolism (VTE spectrum) from the lung injury, which is the main target organ of SARS-CoV-2.

We did not find any association between platelet count and VTE, confirming findings that coagulopathy in COVID-19 is a distinct entity compared to disseminated intravascular coagulation (DIC).(40) The thrombosis phenomena in COVID-19 occurred on an earlier phase, thus do not reflect the state of consumptive coagulopathy.(7,9,41,42)

Finally, we can not confirm other analyzed variables that were and were not associated with COVID-19 due to only including two studies. Thus, more prospective studies are needed.

## Limitations

This systematic review and meta-analysis have several limitations. First, although the total subjects pooled from eight studies were adequate (n= 1237 subjects), it is worth noting that the reported VTE phenomena are not homogeneous. Two studies reported VTE, four studies reported PE only, and two studies reported DVT only. Second, not all of the studies reported the outcome of interest (mortality, ICU admission, and mechanical ventilation). Thus, we can only perform a subgroup analysis for ICU admission. Third, due to many missing data from included studies, more studies are needed to confirm meta-analysis findings that consisted of two studies. Thus, we can not fully elucidate the representative clinical characteristics associated with VTE in COVID-19 patients. Lastly, most of the studies were from western countries, so findings may not be generalizable for other populations. Finally, the included studies were predominantly retrospective design.

## Conclusions

Venous thromboembolism in patients with COVID-19 contributed significantly to higher in-hospital mortality, ICU admission and the need of mechanical ventilation. Additionally, male gender, older age, as well as higher levels of WBC count, D-Dimer, and LDH were associated with increased risks of developing VTE in COVID-19.

## Data Availability

Data is available upon request

## ACKNOWLEDGEMENT

The authors are grateful to Quinta Febryani Handoyono, MD, for her assistance in figure preparation.

## AUTHORS CONTRIBUTION

JH took full responsibility for the project, including creating the concept, designing the study, data accuracy, as well as writing the whole manuscript. JH and ICSP acquired the data. JH, ICSP, HFHD, IC performed extensive research on the topic. All authors contributed to manuscript writing. AC dan LPS review and revised the manuscript. HFHD and IC completed data conversion. JH performed the statistical analysis. All authors approve this manuscript.

## REFERENCES

1. World Health Organization. Coronavirus disease (COVID-19) Situation Report – 132 [Internet]. [cited 2020 Jun 1]. Available from: https://www.who.int/docs/default-source/coronaviruse/situation-reports/20200531-covid-19-sitrep-132.pdf?sfvrsn=d9c2eaef_2

2. Haematology TL. COVID-19 coagulopathy: an evolving story. Lancet Haematol. 2020 Jun 1;7(6):e425.

3. Zhang Y, Xiao M, Zhang S, Xia P, Cao W, Jiang W, et al. Coagulopathy and Antiphospholipid Antibodies in Patients with Covid-19. N Engl J Med. 2020 Apr 23;382(17):e38.

4. Tang N, Bai H, Chen X, Gong J, Li D, Sun Z. Anticoagulant treatment is associated with decreased mortality in severe coronavirus disease 2019 patients with coagulopathy. J Thromb Haemost. 2020;18(5):1094–9.

5. Tang N, Li D, Wang X, Sun Z. Abnormal coagulation parameters are associated with poor prognosis in patients with novel coronavirus pneumonia. J Thromb Haemost. 2020 Apr 1;18(4):844–7.

6. Fox SE, Akmatbekov A, Harbert JL, Li G, Brown JQ, Vander Heide RS. Pulmonary and Cardiac Pathology in Covid-19: The First Autopsy Series from New Orleans [Internet]. Pathology; 2020 Apr [cited 2020 Jun 8]. Available from: http://medrxiv.org/lookup/doi/10.1101/2020.04.06.20050575

7. McGonagle D, O’Donnell JS, Sharif K, Emery P, Bridgewood C. Immune mechanisms of pulmonary intravascular coagulopathy in COVID-19 pneumonia. Lancet Rheumatol [Internet]. 2020 May [cited 2020 Jun 5]; Available from: https://linkinghub.elsevier.com/retrieve/pii/S2665991320301211

8. Leisman DE, Deutschman CS, Legrand M. Facing COVID-19 in the ICU: vascular dysfunction, thrombosis, and dysregulated inflammation. Intensive Care Med [Internet]. 2020 Apr 28 [cited 2020 May 4]; Available from: http://link.springer.com/10.1007/s00134-020-06059-6

9. Cattaneo M, Bertinato EM, Birocchi S, Brizio C, Malavolta D, Manzoni M, et al. Pulmonary Embolism or Pulmonary Thrombosis in COVID-19? Is the Recommendation to Use High-Dose Heparin for Thromboprophylaxis Justified? Thromb Haemost [Internet]. 2020 Apr 29 [cited 2020 Jun 8]; Available from: http://www.thieme-connect.de/DOI/DOI?10.1055/s-0040-1712097

10. Klok FA, Kruip Mjha, van der Meer Njm, Arbous MS, Gommers Dampj, Kant KM, et al. Incidence of thrombotic complications in critically ill ICU patients with COVID-Thromb Res [Internet]. 2020 Apr 10 [cited 2020 May 27]; Available from: http://www.sciencedirect.com/science/article/pii/S0049384820301201

11. Cui S, Chen S, Li X, Liu S, Wang F. Prevalence of venous thromboembolism in patients with severe novel coronavirus pneumonia. Journal of Thrombosis and Haemostasis [Internet]. [cited 2020 May 27];n/a. Available from: https://onlinelibrary.wiley.com/doi/abs/10.1111/jth.14830

12. Wan X, Wang W, Liu J, Tong T. Estimating the sample mean and standard deviation from the sample size, median, range and/or interquartile range. BMC Med Res Methodol [Internet]. 2014 Dec [cited 2020 Jun 8];14(1). Available from: http://bmcmedresmethodol.biomedcentral.com/articles/10.1186/1471-2288-14-135

13. Artifoni M, Danic G, Gautier G, Gicquel P, Boutoille D, Raffi F, et al. Systematic assessment of venous thromboembolism in COVID-19 patients receiving thromboprophylaxis: incidence and role of D-dimer as predictive factors. J Thromb Thrombolysis [Internet]. 2020 May 25 [cited 2020 May 27]; Available from: http://link.springer.com/10.1007/s11239-020-02146-z

14. Leonard-Lorant I, Delabranche X, Severac F, Helms J, Pauzet C, Collange O, et al. Acute Pulmonary Embolism in COVID-19 Patients on CT Angiography and Relationship to D-Dimer Levels. Radiology. 2020 Apr 23;201561.

15. Poyiadji N, Cormier P, Patel PY, Hadied MO, Bhargava P, Khanna K, et al. Acute Pulmonary Embolism and COVID-19. Radiology. 2020 May 14;201955.

16. Grillet F, Behr J, Calame P, Aubry S, Delabrousse E. Acute Pulmonary Embolism Associated with COVID-19 Pneumonia Detected by Pulmonary CT Angiography. Radiology. 2020 Apr 23;201544.

17. Demelo-Rodríguez P, Cervilla-Muñoz E, Ordieres-Ortega L, Parra-Virto A, Toledano-Macías M, Toledo-Samaniego N, et al. Incidence of asymptomatic deep vein thrombosis in patients with COVID-19 pneumonia and elevated D-dimer levels. Thromb Res. 2020 Aug 1;192:23–6.

18. Zhang Li, Feng Xiaokai, Zhang Danqing, Jiang Chunguo, Mei Heng, Wang Jing, et al. Deep Vein Thrombosis in Hospitalized Patients with Coronavirus Disease 2019 (COVID-19) in Wuhan, China: Prevalence, Risk Factors, and Outcome. Circulation [Internet]. [cited 2020 May 27];0(0). Available from: https://www.ahajournals.org/doi/10.1161/CIRCULATIONAHA.120.046702

19. Middeldorp S, Coppens M, Haaps TF van, Foppen M, Vlaar AP, MCA Müller, et al. Incidence of venous thromboembolism in hospitalized patients with COVID-19. J Thromb Haemost [Internet]. [cited 2020 May 27];n/a(n/a). Available from: https://onlinelibrary.wiley.com/doi/abs/10.1111/jth.14888

20. Bompard F, Monnier H, Saab I, Tordjman M, Abdoul H, Fournier L, et al. Pulmonary embolism in patients with Covid-19 pneumonia. Eur Respir J [Internet]. 2020 Jan 1 [cited 2020 May 27]; Available from: https://erj.ersjournals.com/content/early/2020/05/07/13993003.01365-2020

21. 10.4.3.1 Recommendations on testing for funnel plot asymmetry [Internet]. [cited 2020 Jun 8]. Available from: https://handbook-5-1.cochrane.org/chapter_10/10_4_3_1_recommendations_on_testing_for_funnel_plot_asymmetry.htm

22. Terpos E, Ntanasis□Stathopoulos I, Elalamy I, Kastritis E, Sergentanis TN, Politou M, et al. Hematological findings and complications of COVID [19. Am J Hematol [Internet]. 2020 May 23 [cited 2020 Jun 8]; Available from: https://onlinelibrary.wiley.com/doi/abs/10.1002/ajh.25829

23. Fatal Pulmonary Thromboembolism in SARS-CoV-2-Infection. Cardiovasc Pathol. 2020;48:107227.

24. Thachil J, Tang N, Gando S, Falanga A, Cattaneo M, Levi M, et al. ISTH interim guidance on recognition and management of coagulopathy in COVID-19. J Thromb Haemost. 2020;18(5):1023–6.

25. Esmon CT. Basic Mechanisms and Pathogenesis of Venous Thrombosis. Blood Rev. 2009 Sep;23(5):225–9.

26. Hoffmann M, Kleine-Weber H, Schroeder S, Krüger N, Herrler T, Erichsen S, et al. SARS-CoV-2 Cell Entry Depends on ACE2 and TMPRSS2 and Is Blocked by a Clinically Proven Protease Inhibitor. Cell [Internet]. 2020 Mar 5 [cited 2020 Apr 13];0(0). Available from: https://www.cell.com/cell/abstract/S0092-8674(20)30229-4

27. Astuti I, Ysrafil. Severe Acute Respiratory Syndrome Coronavirus 2 (SARS-CoV-2): An overview of viral structure and host response. Diabetes Metab Syndr [Internet]. 2020 Apr 18 [cited 2020 May 14]; Available from: https://www.ncbi.nlm.nih.gov/pmc/articles/PMC7165108/

28. Yan Shi-Fang, Mackman Nigel, Kisiel Walter, Stern David M., Pinsky David J. Hypoxia/Hypoxemia-Induced Activation of the Procoagulant Pathways and the Pathogenesis of Ischemia-Associated Thrombosis. Arterioscler Thromb Vasc Biol. 1999 Sep 1;19(9):2029–35.

29. Levi M, van der Poll T. Inflammation and coagulation. Crit Care Med. 2010 Feb;38(2 Suppl):S26–34.

30. Pal R, Bhansali A. COVID-19, diabetes mellitus and ACE2: The conundrum. Diabetes Res Clin Pract [Internet]. 2020 Apr 1 [cited 2020 Jun 10];162. Available from: https://www.diabetesresearchclinicalpractice.com/article/S0168-8227(20)30382-X/abstract

31. Rizzo P, Vieceli Dalla Sega F, Fortini F, Marracino L, Rapezzi C, Ferrari R. COVID-19 in the heart and the lungs: could we “Notch” the inflammatory storm? Basic Res Cardiol [Internet]. 2020 [cited 2020 Jun 10];115(3). Available from: https://www.ncbi.nlm.nih.gov/pmc/articles/PMC7144545/

32. Wang X, Khaidakov M, Ding Z, Mitra S, Lu J, Liu S, et al. Cross-talk between inflammation and angiotensin II: studies based on direct transfection of cardiomyocytes with AT1R and AT2R cDNA. Exp Biol Med Maywood NJ. 2012 Dec;237(12):1394–401.

33. Varga Z, Flammer AJ, Steiger P, Haberecker M, Andermatt R, Zinkernagel AS, et al. Endothelial cell infection and endotheliitis in COVID-19. The Lancet. 2020 May 2;395(10234):1417–8.

34. Ackermann M, Verleden SE, Kuehnel M, Haverich A, Welte T, Laenger F, et al. Pulmonary Vascular Endothelialitis, Thrombosis, and Angiogenesis in Covid-19. N Engl J Med. 2020 May 21;0(0):ull.

35. Wang T, Chen R, Liu C, Liang W, Guan W, Tang R, et al. Attention should be paid to venous thromboembolism prophylaxis in the management of COVID-19. Lancet Haematol. 2020 May;7(5):e362–3.

36. Roach REJ, Cannegieter SC, Lijfering WM. Differential risks in men and women for first and recurrent venous thrombosis: the role of genes and environment. J Thromb Haemost. 2014;12(10):1593–600.

37. Silverstein MD, Heit JA, Mohr DN, Petterson TM, O’Fallon WM, Melton LJ. Trends in the Incidence of Deep Vein Thrombosis and Pulmonary Embolism: A 25-Year Population-Based Study. Arch Intern Med. 1998 Mar 23;158(6):585–93.

38. Tian W, Jiang W, Yao J, Nicholson CJ, Li RH, Sigurslid HH, et al. Predictors of mortality in hospitalized COVID□19 patients: A systematic review and meta□analysis. J Med Virol [Internet]. 2020 May 22 [cited 2020 Jun 8]; Available from: https://onlinelibrary.wiley.com/doi/abs/10.1002/jmv.26050

39. AlGhatrif M, Cingolani O, Lakatta EG. The Dilemma of Coronavirus Disease 2019, Aging, and Cardiovascular Disease: Insights From Cardiovascular Aging Science. JAMA Cardiol [Internet]. 2020 Apr 3 [cited 2020 Jun 11]; Available from: https://jamanetwork.com/journals/jamacardiology/fullarticle/2764300

40. Panigada M, Bottino N, Tagliabue P, Grasselli G, Novembrino C, Chantarangkul V, et al. Hypercoagulability of COVID-19 patients in Intensive Care Unit. A Report of Thromboelastography Findings and other Parameters of Hemostasis. J Thromb Haemost JTH. 2020 Apr 17;

41. Buja LM, Wolf DA, Zhao B, Akkanti B, McDonald M, Lelenwa L, et al. The emerging spectrum of cardiopulmonary pathology of the coronavirus disease 2019 (COVID-19): Report of 3 autopsies from Houston, Texas, and review of autopsy findings from other United States cities. Cardiovasc Pathol. 2020 Sep;48:107233.

42. Belen-Apak FB, Sarıalioğlu F. Pulmonary intravascular coagulation in COVID-19: possible pathogenesis and recommendations on anticoagulant/thrombolytic therapy. J Thromb Thrombolysis [Internet]. 2020 May 5 [cited 2020 Jun 5]; Available from: https://doi.org/10.1007/s11239-020-02129-0

